# Patients’ Experience and Satisfaction towards Virtual Health Care during the COVID-19 Pandemic in Southern Region of Saudi Arabia

**DOI:** 10.1101/2024.08.28.24312750

**Authors:** Ayed A. Shati, Hasan S. Alamri, Abdulaziz M. Al-Garni, Syed E. Mahmood, Awad S. Alsamghan

## Abstract

**Background:** Healthcare providers can use these virtual platforms for delivering medical advice and prescriptions of medications to patients. This study was aimed to explore the patient’s experiences and level of satisfaction regarding virtual health care received during the COVID-19 pandemic. This study also assessed the before and during lockdown sleep quality in these participants.

**Design and Methods:** The current study included 522 participants from Saudi Arabia. Virtual healthcare satisfaction questionnaire was implemented to record the data on patient experience toward the virtual health care during COVID-19 pandemic.

**Results:** Patients’ opinions regarding virtual health care received during the COVID-19 pandemic era are summarized using the “five-point Likert scale”. The mean score of all the five statements is 4.15, reflecting 83% level of satisfaction of participants towards virtual health care. Among participants, mostly agreed that they were able to communicate adequately with doctors, the picture and sound quality of the virtual appointment were good, their privacy was respected during the consultation, they were comfortable during history taking and exams that were done, and the doctor explained solutions including prescribing medicine and/or providing advice. However, there was no significant association found between the type of specialty attended or patient demographic factors and level of satisfaction. Results of subjective sleep parameters assessed before and during lock down are depicted. Results of t-test showed that the mean scores of all the components namely sleep duration (P<0.001); sleep disturbances (P<0.001), sleep latency (P<0.001), daytime dysfunction (P<0.001), habitual sleep efficiency (P<0.001), and subjective sleep quality (P<0.001) significantly differed between two assessment levels. Mean scores of all the components shifted towards higher values, representing worsening of sleep quality during the COVID-19 pandemic lock down.

**Conclusions:** Findings support and conclude that virtual health care system can be adopted in clinical settings.

## Introduction

In face of an exponential increase in cases of COVID-19, healthcare systems worldwide are racing to adopt virtual consultations and treatment approaches [1]. This obviates the need for physical meetings between patients and healthcare providers, therefore increasing the safety for patients as well as for healthcare providers. The virtual or digital healthcare is cost effective and secure use of data and communication technologies to deal with several medical conditions without seeing the patient face to face [2]. Healthcare providers can use different virtual platforms such as real-time audio-video meeting or audio/text messaging to address patient’s concerns. Healthcare providers can use these virtual platforms for delivering medical advice and prescriptions of medications to patients [1,3]. The use of virtual health care has been suggested as a method to maintain a continuum of healthcare for patients [4,5]. As a result, the prevalence of virtual care has rapidly increased during this COVID-19 pandemic. There has been exponential increase in virtual healthcare consultations in the United States, China, Canada, UK, Italy, Germany, India, Cameroon, Ethiopia, Kenya, Malawi, Mozambique, South Africa, South Sudan, and Uganda during this COVID-19 [1]. In 2020, one of the study conducted in USA reported that the virtual consultations for telemedicine increased within four weeks from less than 1% to 70% [6]. In addition to these findings, several studies documented that virtual health care system along with telemedicine contribute to various health-related aspects such as to reduce crowding in hospitals and clinics, preventing non-emergency cases from unnecessary exposure to COVID-19 zones, and providing safety for both patients and healthcare providers [2,7,8,9]. However, there are certain concerns regarding the use of virtual healthcare system such as only little percentage of physicians and patients are adequately educated on how to utilize these digital services [2]. Other major concerns are the common problems in connectivity and how much the patients are satisfied with this virtual health care system. A study was conducted in 2019 about assessment of knowledge, perception and willingness of virtual consultation and telemedicine among physicians [10]. Authors reported that the main problems associated with virtual consultations and telemedicine was patient’s privacy, high cost of equipment, lack of training and lack of connection between information technology expert and clinicians.

With regard to patient’s level of satisfaction for virtual healthcare system, one retrospective cohort study was conducted in New York City to measure the patient satisfaction with telemedicine during the COVID-19 pandemic [11]. This study reported that patient satisfaction with virtual consultation (video visits) is high. Another pilot study was carried out to measure the patient satisfaction with telemedicine in home health services for the Older person/persons [12]. Authors reported that telemedicine was significantly effective in reducing the average number of clinic visits per month. Overall 72% of patients were satisfied with telemedicine.

Digital health started in Saudi Arabia since 2010. In 2017, the Ministry of Health launched the Tele-Health application called “Seha”. This application is used all over the kingdom for virtual medical consultation through voice and video calls. Since the surge of COVID-19 cases started, the Ministry of Health of Saudi Arabia suspended clinical visit, canceled nonemergency appointments and encouraged the community to use digital applications either for counseling or to attend appointments remotely in order to control the spread of disease. In order to measure the level of satisfaction of patients for virtual consultation during COVID-19, a study was conducted that showed that patients were highly satisfied. The level of satisfaction was 80.4% [13]. This study has a sample size limitation. However, overall there is a scarcity of related data that can signify the patient’s satisfaction level for the virtual health care system.

Above discussed literature signifies the importance of virtual healthcare system during this COVID-19 pandemic. But there are some important concerns such as the level of satisfaction of patients for this virtual health care system. There is scarcity of such reports from the region of Saudi Arabia. Further studies are required to assess patient perceptions of receiving medical care virtually, its advantages and challenges during the pandemic. Thus, current study was aimed to evaluate the patient experience toward the virtual health care during the COVID-19 pandemic in Saudi Arabia.

## Design and methods

### Study design and Participants

The current study is an observational descriptive cross-sectional study. The aim of the study was to explore patients’ opinions regarding the virtual health care received during the COVID-19 pandemic, to associate the type of specialized medical services received and patient sociodemographic factors with patients’ level of satisfaction towards the virtual health care received, to explore the factors associated with dissatisfaction among patients during the virtual healthcare sessions, and to compare the before lock down and during lock down sleep quality to explore the impact of COVID-19 pandemic. The study was conducted from August 29, 2020 to November 29, 2020. Initially we recruited 583 participants from Saudi Arabia who have used virtual healthcare platforms at least once during the pandemic and aged ≥18 years. Among these participants, 61 participants were dropped out due to missing data, health related issues, lack of technical ability, and currently not residing in Saudi Arabia. Finally, 522 participants were included in this study. We hypothesized that the patients prefer and are satisfied regarding virtual health care during the COVID-19 pandemic in Saudi Arabia. The research question was: What are the patients’ experiences and satisfaction regarding virtual health care during the COVID-19 pandemic in Saudi Arabia?

### Participant inclusion criteria

Participants should have access to required technology and the internet, is comfortable with using audio/video technology, has no physical/sensory/cognitive disabilities that would limit audio/video conference use, has access to email/chat, and is comfortable with using the modality.

### Assessment of patient experience toward the virtual health care

Virtual healthcare satisfaction questionnaire was implemented to record the data on patient experience toward the virtual health care. The questionnaire’s purpose is to evaluate the satisfaction and viewpoints of patients regarding digital healthcare during COVID-19. This questionnaire contains questions regarding sociodemographic characteristics (Table 1) and telemedicine satisfaction (Supplementary Table 1).

**Table 1.**
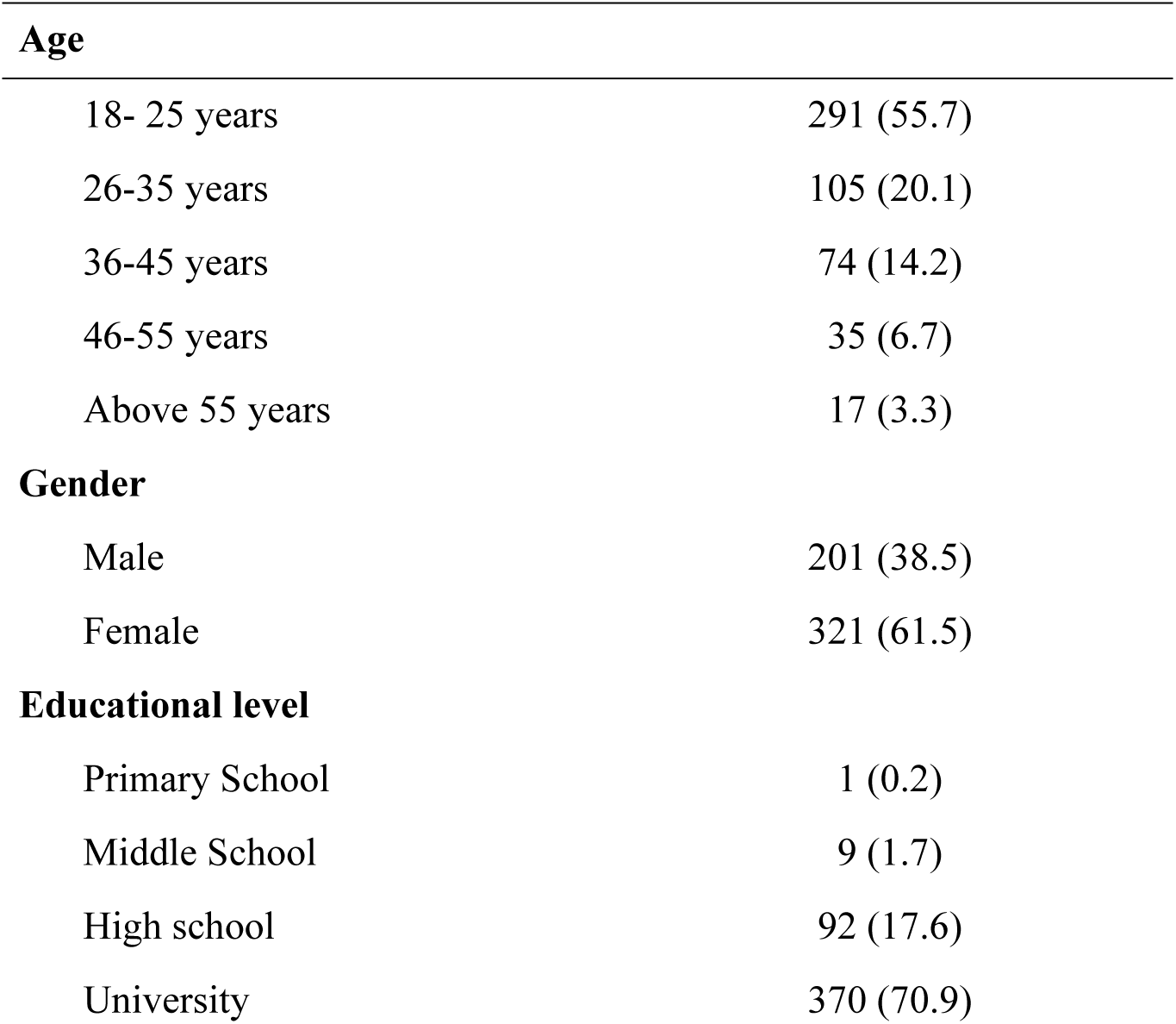
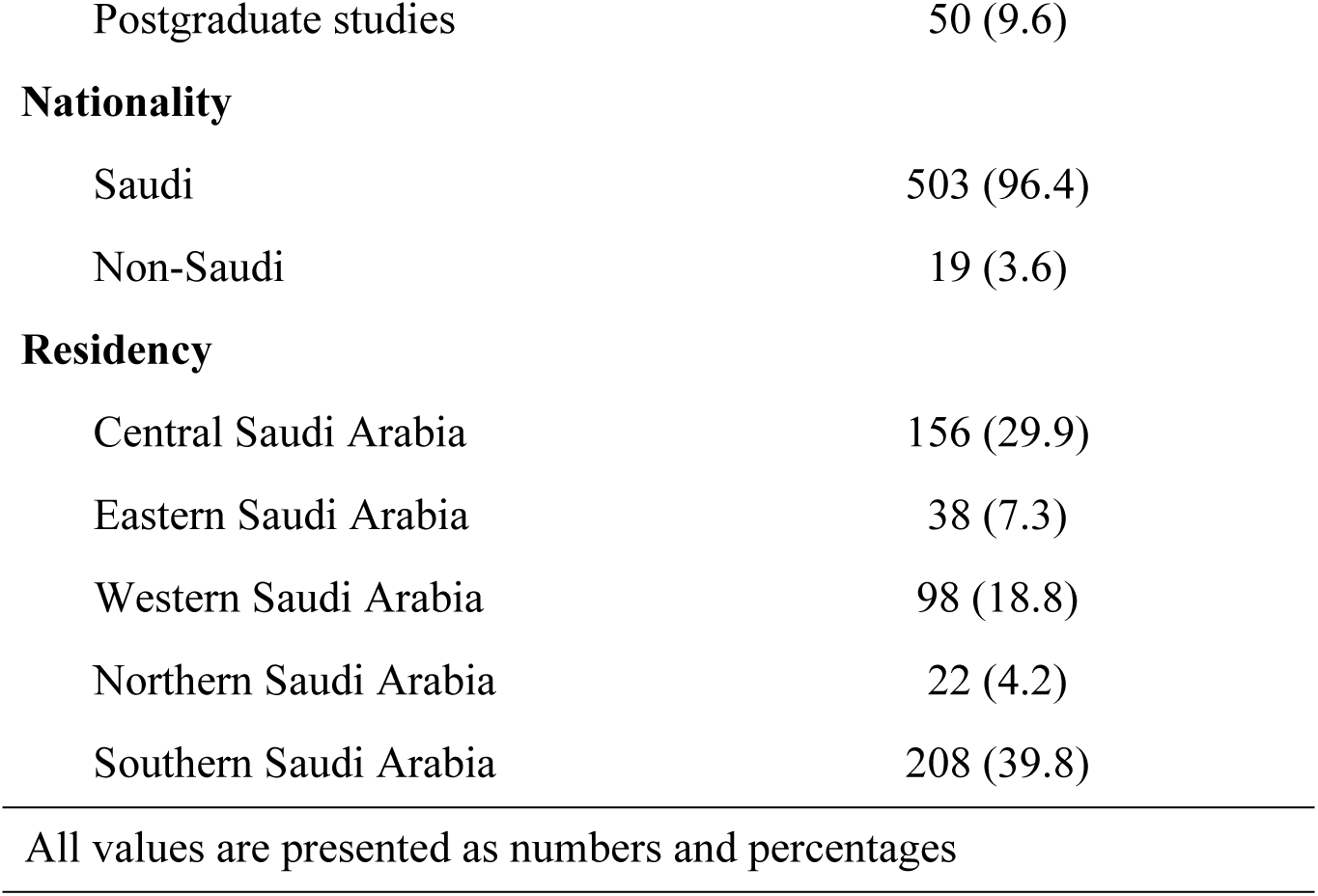
Demographics characteristics of the participants (n=522) Age.

| Age |  |
| --- | --- |
| 18- 25 years | 291 (55.7) |
| 26-35 years | 105 (20.1) |
| 36-45 years | 74 (14.2) |
| 46-55 years | 35 (6.7) |
| Above 55 years | 17 (3.3) |
| Gender |  |
| Male | 201 (38.5) |
| Female | 321 (61.5) |
| Educational level |  |
| Primary School | 1 (0.2) |
| Middle School | 9 (1.7) |
| High school | 92 (17.6) |
| University | 370 (70.9) |
| Postgraduate studies | 50 (9.6) |
| **Nationality** |  |
| Saudi | 503 (96.4) |
| Non-Saudi | 19 (3.6) |
| **Residency** |  |
| Central Saudi Arabia | 156 (29.9) |
| Eastern Saudi Arabia | 38 (7.3) |
| Western Saudi Arabia | 98 (18.8) |
| Northern Saudi Arabia | 22 (4.2) |
| Southern Saudi Arabia | 208 (39.8) |

All values are presented as numbers and percentages

### Ethical approval

Current study was approved (Approval No.: ECM#2020-1002) by the Research Ethics Committee, King Khalid University, Saudi Arabia on 19/08/2020. All participants provided their oral and written consent. Participants were assured that the information will be handled anonymously and for scientific purposes only.

### Statistical Analysis

The data analysis related to evaluation of patient experience toward the virtual health care included two stages. The first stage included a descriptive analysis, in which numerical variables were reported in terms of means, standard deviations and standard errors. The second stage included hypothesis testing using the Pearson Chi-Square test and Likert scale analysis. Moreover, we employed paired samples t-test to compare the responses of participants for the before lock down and during lock down sleep quality. All the data was analyzed in SPSS (Version 22.0).

## Results

### Descriptive Analysis

#### Socio-demographic information

Socio-demographic information of all the participants is presented in Table 1. Among the participants, 291 (55.7%) belonged to younger age group of 18-25 years and 80.5% of total participants had received university level education. Majority of the participants (n=503; 96.4%) included in the current study are Saudi nationals.

#### Patients’ opinions regarding virtual health care and Likert Scale Analysis

Patients’ opinions regarding virtual health care received during the COVID-19 pandemic era are summarized in Supplementary Table 1. The “five-point Likert scale” gives weight as value 4.21 – 5.00 (Strongly Agree), 3.41 – 4.20 (Agree), 2.61 – 3.40 (Neutral), 1.81 – 2.60 (Disagree), and 1.00 – 1.80 (Strongly Disagree). cis presented in Table 2. Results showed that most participants agreed that they were able to communicate adequately with doctors (189 (36.2%) strongly agreed and 212 (40.6%) agreed), the picture and sound quality of the virtual appointment were good (203 (38.9) strongly agreed and 214 (41%) agreed), their privacy was respected during the consultation (267 (51.1) strongly agreed and 196 (37.5) agreed), they were comfortable during history taking and exams that were done (210 (40.2) strongly agreed and 218 (41.8) agreed), and the doctor explained solutions including prescribing medicine and/or providing advice (222 (42.5) strongly agreed and 196 (37.5) agreed). The mean and standard deviations of all the five statements are presented in Table 3. Overall, statement 3 (My privacy was respected during consultations) has the highest satisfaction level of 4.36 (Strongly agree). The mean score of all the five statements is 4.15, reflecting 83% level of satisfaction of participants towards virtual health care (Table 3).

**Table 2.** Patients’ level of satisfaction towards the virtual health care received (5-point Likert scale analysis)

| S.No. | Statement | Strongly Agree<br>n (%) | Agree<br>n (%) | Neutral<br>n (%) | Disagree<br>n (%) | Strongly Disagree<br>n (%) |
| --- | --- | --- | --- | --- | --- | --- |
| 1. | I was able to communicate adequately with my doctor | 189 (36.2) | 212 (40.6) | 85 (16.3) | 24 (4.6) | 12 (2.3) |
| 2. | The picture and sound quality were good | 203 (38.9) | 214 (41) | 78 (14.9) | 17 (3.3) | 10 (1.9) |
| 3. | My privacy was respected during consultations | 267 (51.1) | 196 (37.5) | 45 (8.6) | 7 (1.3) | 7 (1.3) |
| 4. | I was comfortable during history taking and exams that were done | 210 (40.2) | 218 (41.8) | 65 (12.5) | 15 (2.9) | 14 (2.7) |
| 5. | The doctor explained to me solutions including prescribing medicine and/or providing advice | 222 (42.5) | 196 (37.5) | 63 (12.1) | 22 (4.2) | 19 (3.6) |

**Table 3.**
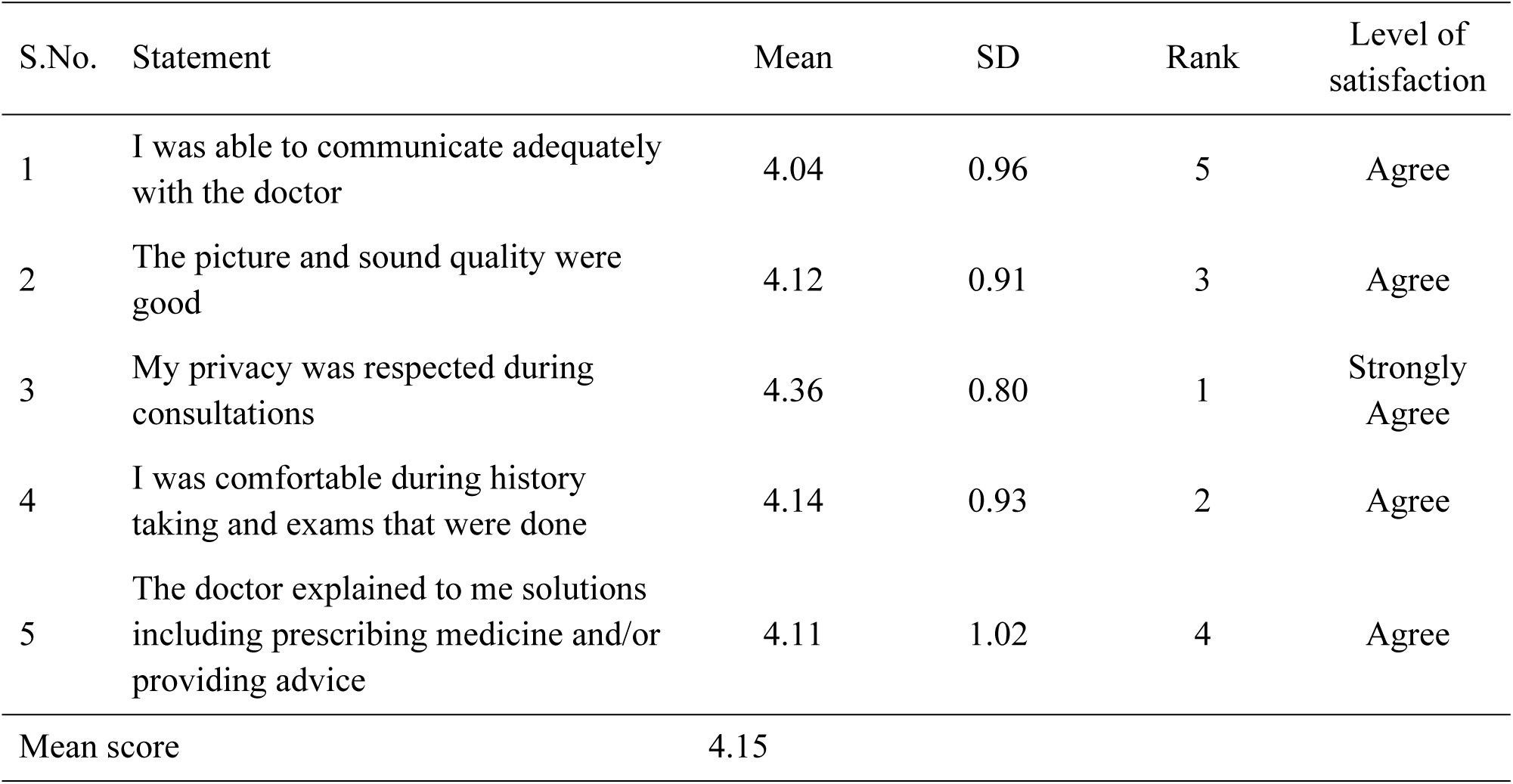
Patients’ level of satisfaction towards the virtual health care received (5-point Likert scale analysis)

| S.No. | Statement | Mean | SD | Rank | Level of satisfaction |
| --- | --- | --- | --- | --- | --- |
| 1 | I was able to communicate adequately with the doctor | 4.04 | 0.96 | 5 | Agree |
| 2 | The picture and sound quality were good | 4.12 | 0.91 | 3 | Agree |
| 3 | My privacy was respected during consultations | 4.36 | 0.80 | 1 | Strongly Agree |
| 4 | I was comfortable during history taking and exams that were done | 4.14 | 0.93 | 2 | Agree |
| 5 | The doctor explained to me solutions including prescribing medicine and/or providing advice | 4.11 | 1.02 | 4 | Agree |
| Mean score |  | 4.15 |  |  |  |

### Bivariate Analysis

#### Testing the association between the types of specialized medical services received with the patients’ level of satisfaction towards the virtual health care received (Chi Square test)

Table 4 present the association between the types of specialized medical services received and the patient’s level of satisfaction towards the virtual health care (reported using the Pearson chisquare test). Results showed no significant association between the type of specialized medical services received and the patient’s level of satisfaction towards the virtual health care.

**Table 4.** Testing the association between the types of specialized medical services received with the patients’ level of satisfaction towards the virtual health care received (Chi Square test).

|  | Level of satisfaction |  |  |  |  |  |
| --- | --- | --- | --- | --- | --- | --- |
| Specialty attended | Strongly Agree<br>n (%) | Agree<br>n (%) | Neutral<br>n (%) | Disagree<br>n (%) | Strongly Disagree<br>n (%) | P-value |
| Internal medicine | 34 (6.51) | 49 (9.38) | 9 (1.72) | 5 (0.95) | 2 (0.38) | 0.223 |
| General surgery | 10 (1.91) | 11 (2.10) | 0 (0) | 0 (0) | 0 (0) |  |
| Obstetrics and gynecology | 8 (1.53) | 5 (0.95) | 2 (0.38) | 0 (0) | 0 (0) |  |
| Pediatrics | 15 (2.87) | 17 (3.25) | 2 (0.38) | 0 (0) | 0 (0) |  |
| Family medicine | 33 (6.32) | 26 (4.98) | 5 (0.95) | 2 (0.38) | 2 (0.38) |  |
| Orthopedic | 8 (1.53) | 9 (1.72) | 2 (0.38) | 2 (0.38) | 0 (0) |  |
| Dental | 12 (2.29) | 12 (2.29) | 1 (0.19) | 1 (0.19) | 1 (0.19) |  |
| Psychiatry | 8 (1.53) | 3 (0.57) | 0 (0) | 1 (0.19) | 1 (0.19) |  |
| General | 51 (9.77) | 50 (9.57) | 13 (2.49) | 0 (0) | 7 (1.34) |  |
| Ophthalmology | 14 | 11 (2.10) | 2 (0.38) | 0 (0) | 0 (0) |  |
| Others | 32 (6.13) | 30 (5.74) | 13 (2.49) | 0 (0) | 1 (0.19) |  |
| No significant association was found between type of specialized medical services received and patient's level of satisfaction at 0.05 level of significance. |  |  |  |  |  |  |

#### Testing the association between socio-demographic factors with patients’ level of satisfaction towards the virtual health care received (Chi Square test)

Table 5 presents the association between patient’s demographic factors and their level of satisfaction towards the virtual health care (reported using the Pearson chi-square). No significant association was found between the demographic factors and patient level of satisfaction towards the virtual health care.

**Table 5.** Testing the association between socio-demographic factors with patients’ level of satisfaction towards the virtual health care received (Chi Square test).

| Level of satisfaction |  |  |  |  |  |  |
| --- | --- | --- | --- | --- | --- | --- |
| Demographics | Strongly Agree<br>n (%) | Agree<br>n (%) | Neutral<br>n (%) | Disagree<br>n (%) | Strongly Disagree<br>n (%) | P-value |
| Age |  |  |  |  |  |  |
| 18-25 years | 129 (24.71) | 126 (24.13) | 26 (4.98) | 3 (0.57) | 7 (1.34) | 0.159 |
| 26-35 years | 40 (7.66) | 46 (8.81) | 12 (2.29) | 5 (0.95) | 2 (0.38) |  |
| 36-45 years | 31 (5.93) | 33 (6.32) | 6 (1.14) | 2 (0.38) | 2 (0.38) |  |
| 46-55 years | 12 (2.29) | 14 (2.68) | 5 (0.95) | 1 (0.19) | 3 (0.57) |  |
| Above 55 years | 13 (2.49) | 4 (0.76)) | 0 (0) | 0 (0) | 0 (0) |  |
| Gender |  |  |  |  |  |  |
| Male | 74 (14.17) | 93 (17.81) | 21 (4.02) | 5 (0.95) | 8 (1.53) | 0.157 |
| Female | 151 (28.92) | 130 (24.90) | 28 (5.36) | 6 (1.14) | 6 (1.14) |  |
| Educational level |  |  |  |  |  |  |
| Primary School | 1 (0.19) | 0 (0) | 0 (0) | 0 (0) | 0 (0) | 0.095 |
| Middle School | 2 (0.38) | 3 (0.57) | 2 (0.38) | 0 (0) | 2 (0.38) |  |
| High school | 46 (8.81) | 39 (7.47) | 5 (0.95) | 1 (0.19) | 1 (0.19) |  |
| University | 153 (29.31) | 161 (30.84) | 39 (7.47) | 8 (1.53) | 9 (1.72) |  |
| Postgraduate studies | 23 (4.40) | 20 (3.83) | 3 (0.57) | 2 (0.38) | 2 (0.38) |  |
| Nationality |  |  |  |  |  |  |
| Saudi | 215 (41.18) | 217 (41.57) | 47 (9.00) | 10 (1.91) | 14 (2.68) | 0.649 |
| Non-Saudi | 10 (1.91) | 6 (1.14) | 2 (0.38) | 1 (0.19) | 0 (0) |  |
| No association found between all demographic factors and patient level of satisfaction at the 0.05 level of significance. |  |  |  |  |  |  |

#### Exploring the factors associated with dissatisfaction among patients during the virtual health care sessions

We also explored the dissatisfaction among patients during their virtual healthcare sessions (Table 6). The dissatisfaction includes problem related with internet connection, taking long time and difficulty in explaining different concern.

**Table 6.** Exploring the factors associated with dissatisfaction among patients during the virtual healthcare sessions.

| Problems encountered | Overall experience satisfaction |  |  |  |  |
| --- | --- | --- | --- | --- | --- |
|  | Very Satisfied<br>n (%) | Satisfied<br>n (%) | Neutral<br>n (%) | Dissatisfied<br>n (%) | Very Dissatisfied<br>n (%) |
| Internet connection | 16 (3.06) | 37 (7.08) | 30 (5.74) | 5 (0.95) | 10 (1.91) |
| Difficulty in using the platform | 7 (1.34) | 16 (3.06) | 13 (2.49) | 3 (0.57) | 5 (0.95) |
| Long time until start | 32 (6.13) | 54 (10.34) | 66 (12.64) | 5 (0.95) | 10 (1.91) |
| Difficulty in clarifying my concerns | 27 (5.17) | 42 (8.04) | 45 (8.62) | 9 (1.72) | 5 (0.95) |
| Healthcare provider was uncooperative | 0 (0) | 2 (0.38) | 14 (2.68) | 2 (0.38) | 6 (1.14) |
| None | 107 (20.49) | 74 (14.17) | 20 (3.83) | 0 (0) | 4 (0.76) |

### Analyses of the sleep quality before and during COVID-19 lock down

#### Frequency distribution of participants with good and bad sleep before and during COVID-19 pandemic lock down

Frequency distribution of good and bad sleep sleepers was computed based on the PSQI global scores obtained at the two assessment levels, i.e., before and during the COVID-19 pandemic lock down. Before lockdown scores showed that 87% participants were good sleepers (i.e., PSQI < 5) and 13% were bad sleepers (i.e., PSQI ≥ 5), whereas during lock down assessment showed that the distribution of bad sleepers increased significantly increased (p<0.001) from 13% to 83% and the distribution of good sleepers significantly reduced (p<0.001) from 87% to 17%.

#### Subjective sleep parameters

Results of subjective sleep parameters assessed before and during lock down are depicted in Table 7. Results of t-test showed that the mean scores of all the components namely sleep duration (P<0.001); sleep disturbances (P<0.001), sleep latency (P<0.001), daytime dysfunction (P<0.001), habitual sleep efficiency (P<0.001), and subjective sleep quality (P<0.001) significantly differed between two assessment levels. Mean scores of all the components shifted towards higher values, representing worsening of sleep quality during the COVID-19 pandemic lock down.

## Discussion

The novel coronavirus disease-19 (COVID-19) pandemic has altered several aspects such as economy, society, and healthcare system. Healthcare systems worldwide are racing to adopt virtualized consultations and treatment approaches [1]. Telehealth programs such as virtualized consultations and treatment approaches overcome physical barriers to provide patients and caregivers access to convenient medical care [6]. As a result, the prevalence of virtual care has rapidly increased thought out the world during this COVID-19 pandemic [1]. However, there are certain concerns regarding the use of virtual healthcare system, reflecting the need of an assessment of level of satisfaction of people for these virtual interface platforms [2]. For example one study was recently conducted to assess the knowledge, perception and willingness for virtual consultation and telemedicine [10]. Authors reported that lack of training and lack of connection between information technology expert and clinicians, lack of training in patients, patient’s care about their privacy, etc were the main problems associated with virtual consultations and telemedicine.

Despite of the facts, one study was previously conducted in Saudi Arabia, where authors evaluated the physician’s willingness towards adopting telemedicine in clinical practice [10]. Authors reported that most of the participants showed positive perceptions of virtual health care and are willing to adopt it in clinical practice. However, there are several major barriers such as privacy issues, lack of training, cost and issues related to information and communication technology for the adoption of virtual health care system.

One study was conducted in British Columbia, Canada regarding the satisfaction of patients about virtual visits and patient centered care [14]. This study reported that about 93.2% of respondents saying their virtual visits were of high quality, 91.2% participants reporting their virtual visits as “very” or “somewhat” helpful to resolve their health issue, and 79% were having confidence in the security and privacy of this virtual healthcare system. This is somewhat in line with findings of our current study, where responses of majority of the participants reflect that they were comfortable and satisfied with virtual health care. These findings together support for the adaptation of virtual health care system in the clinical settings.

In current study the level of satisfaction towards virtual healthcare was high (83%). This echoes the finding of previous study published about patient satisfaction toward a virtual healthcare in endocrinology clinics in Saudi Arabia [13] Authors reported that the level of satisfaction for virtual health care system was about 80.4%. A study was conducted in Chile aimed to assess patient satisfaction with tele-neurology and it was found that the level of satisfaction was high about 97% [15]. One study evaluated usability and satisfaction of telemedicine for head and neck ambulatory visits [16]. This study reported that the patients were highly satisfied with telemedicine. Another study was aimed to characterized telemedicine and correlates it with patient’s level of satisfaction [17]. This study showed that the overall 85% of patients rated their satisfaction with their telemedicine physician five stars on scales of 0 to 5. About the problems participants faced during virtual session, long waited time was on top followed by difficult to clarify their concerns and internet connection problems. Our findings are supported by another recent study in Saudi Arabia and nearly half of the participants wanted to continue using virtual services even after the COVID-19 pandemic was over [18]. More effort should be made to increase patient awareness and knowledge about virtual clinics. Saudi Arabian citizens believe that telemedicine saves time, labour and costs and is an effective tool to treat coronavirus patients at a safe distance. [19]

Our findings are also comparable to the Argentina study which created and implemented a virtual care program for patients with URTI during the epidemiological outbreak. [20]. Based on their work, they deemed this new care strategy could help prevent hospital overcrowding, reduce long delays inattention (or at least offer the possibility of waiting at home), avoid unnecessary referrals, and limit infections in waiting rooms. The inability to meet the health-care professional face-to-face was reported by 53.8% as the most important disadvantage in another cross-sectional study conducted among patients who had experienced virtual clinics in primary healthcare centers in Riyadh, Saudi Arabia [21].

The key advantage of telemedicine is its mobility, and it presented a perfect match in the current public health policy of social distancing and cancellation of non-urgent consultations and surgeries [22]. The virtual clinic services in Saudi Arabia provides a wide range of health-care consultations in different fields such as family medicine, general surgery, ophthalmology, ENT, Obstetrics /Gynecology, dermatology, chronic diseases, pediatrics and vaccinations However, there are no current data on the exact number of virtual clinics. [23]. The Sehhaty mobile application can be used by a patient to directly book an appointment if the service is available in the healthcare facility. The “Seha Virtual Hospital”, supports a total of 130 hospitals around the country and provides specialized virtual clinics such as psychiatry, cardiac, endocrine, and diabetes clinics [24]. Other applications (TETAMMAN, TABAUD, TAWAKKALNA) that have been launched have improved the quality of using telemedicine activities and facilitated all health services [25]. This accelerated digital health innovation in Saudi Arabia and allowed patients to recognise the benefits of digital health. This has huge potential for increasing continuous patient engagement with PHCs [26].

Specific patient education experiences might be most helpful for patients in learning about virtual care. People with lived experience could be resources for educating patients and community-based knowledge sharing was also encouraged (27). Our sample size had less number of older people, the probable reason is they were less likely to have used or familiar telehealth. The results of an Australian population support this need for better support for older people to access remote modes of care (28). Willingness to use telemedicine in future was high in the recipients as well as the providers of healthcare during COVID-19 pandemic in UK(29). The findings from a Saudi study showed that 70.6% of participants were aware of the existence of virtual clinics and 34.6% of participants were post-pandemic users (30). Over 90% Saudis preferred having a virtual appointment over an in-person visit according to a recent study. About half had telephone consultations, while about a third had video calls through hospital-provided platforms; >90% found virtual appointments useful and convenient, and 97.4% were satisfied with their remote consultation experience despite the technical interruptions. (31). Telemedicine has developed rapidly during the COVID-19 pandemic, and its satisfaction measurement factors may be changed in the future. Du, Y. et al developed a scale for evaluating patient satisfaction with telemedicine by applying multidimensional constructs to capture patient satisfaction comprehensively, which involves nine dimensions, such as humanistic care, doctor-patient communication, service efficiency, diagnosis and treatment result, ease of use, system quality, usefulness, privacy and security, overall satisfaction. This scale could be a meaningful tool for future studies (32).

The limitation of this study is the implicit bias usually present in this descriptive, online voluntary response survey, and the relatively low response rate. The dropout rate of participants raises concerns about sample representativeness and potential bias. Moreover, the study’s reliance on voluntary participation introduces self-selection bias, as individuals who are more satisfied or dissatisfied with virtual healthcare may be more inclined to participate. The inclusion criteria focus primarily on technological access and comfort rather than broader factors that could influence patient satisfaction, such as socioeconomic status or health literacy. This narrow focus may limit the generalizability of the findings. Other study limitation is a small sample size and limitation of generalizability of the results due to the small sample size. We hope in the future to have all the required resources to do multicentric /nationwide studies with a larger sample size. However, as the importance of patient satisfaction continues to increase in the provision of health care, this work is considered to be an important step in this direction, particularly in Saudi Arabia.

## Conclusions

Findings from the current study reflected that most of the participants were satisfied with virtual healthcare system. The virtual healthcare could help prevent overcrowding in health facilities especially in such pandemics, decrease delays in attention and avoid unnecessary in-person visits, easing access to healthcare services possibly by providing patients with needed care at appropriate time. All of these could have a significant impact on costs and patient satisfaction but not necessarily displacing in-person visits to hospital and clinics. Future studies with a larger sample size and multicentric in nature need to further explore the patient’s experiences and level of satisfaction regarding virtual health care in Saudi Arabia. This will help to further continue and expand the telehealth services in the Kingdom. It will also give telemedicine providers insights into areas where they can improve their services.

## Data Availability

If the data are all contained within the manuscript and/or Supporting Information files, enter the following: All relevant data are within the manuscript and its Supporting Information files.

## Acknowledgements

The authors are thankful to the Institute of Research and consulting studies at King Khalid University for supporting this research (grant number 4-N-20/21).

## Contributions

All authors contributed significantly & agreed with the manuscript content, have read carefully & approved the final version.

---

**Supplementary Table 1.** Patients’ opinions regarding virtual health care received during the COVID-19 pandemic era.

| Questions | Respondents n (%) |
| --- | --- |
| Who was attending the virtual clinic? (n=522) |  |
| Me | 351 (67.2) |
| Child under my care | 56 (10.7) |
| Older person under my care | 115 (22) |
| How was the virtual clinic conducted? (n=522) |  |
| Sound only | 465 (89.1) |
| Video | 57 (10.9) |
| What was the reason for using a virtual clinic? (n=522) |  |
| Appointment | 95 (18.2) |
| Counseling and medical advice | 349 (66.9) |
| Emergency | 78 (14.9) |
| With what medical specialty did you meet? (n=522) |  |
| Internal medicine | 99 (19) |
| General surgery | 21 (4) |
| Obstetrics and gynecology | 15 (2.9) |
| Pediatrics | 34 (6.5) |
| Family medicine | 68 (13) |
| Orthopedic | 21 (4) |
| Dental | 27 (5.2) |
| Psychiatry | 13 (2.5) |
| General | 121 (23.2) |
| Ophthalmology | 27 (5.2) |
| Others | 76 (14.6) |
| What is your opinion on receiving virtual health care in comparison to traditional, in-person, medical visits? (n=522) |  |
| Better than traditional | 179 (34.3) |
| As good as traditional | 168 (32.2) |
| Worse than traditional | 56 (10.7) |
| Not sure | 119 (22.8) |
| Do you prefer to attend your next appointment/counseling in a non-virtual clinic? (n=522) |  |
| Yes | 236 (45.2) |
| No | 86 (16.5) |
| Maybe | 200 (38.3) |
| Do you prefer to keep using telemedicine platforms after the pandemic ends?<br>(n=522) |  |
| Yes | 217 (41.6) |
| No | 103 (19.7) |
| Maybe | 202 (38.7) |
| Do you feel that the physical presence of a doctor and a physical examination<br>by him/her may give you more trust in the diagnosis / treatment? (n=522) |  |
| Yes | 367 (70.3) |
| No | 35 (6.7) |
| Maybe | 120 (23) |
| Do you think your presence in the clinical health care unit is essential for your<br>confidentiality in treatment?(n=522) |  |
| Yes | 261 (50) |
| No | 108 (20.7) |
| Maybe | 153 (29.3) |
| What problems did you face during these sessions? (n=522) (r=606)* |  |
| Internet connection | 98 (18.8) |
| Difficulty in using the platform | 44 (8.4) |
| Long time until start | 167 (32) |
| Difficulty clarifying my concerns | 128 (24.5) |
| Healthcare provider was uncooperative | 24 (4.6) |
| None | 205 (39.3) |
| Do you think virtual health care helps to reduce the risk of exposure to COVID-19 virus? (n=522) |  |
| Yes | 396 (75.9) |
| No | 30 (5.7) |
| Maybe | 96 (18.4) |
| Overall experience satisfaction(n=522) |  |
| very dissatisfied | 22 (4.2) |
| dissatisfied | 13 (2.5) |
| neutral | 117 (22.4) |
| satisfied | 194 (37.2) |
| very satisfied | 176 (33.7) |

All values are presented as numbers and percentages.
\*n = sample size or total number of cases, r = total number of responses in multiple choices

